# A study protocol for an Observational Feasibility Study on Mass Drug Administration and Serology Integrated with Reactive Case Detection for Vivax Malaria Elimination in Cambodia

**DOI:** 10.64898/2026.02.11.26346129

**Authors:** Soy Ty Kheang, Siv Sovannaroth, Manash Shrestha, Jean Popovici, Ivo Mueller, Leanne J. Robinson, Thao Huynh, Thy Do, Elodie Jambert, Caroline A. Lynch

**Author notes:** Corresponding Author, (MS).

## Abstract

**Background:** *Plasmodium vivax (P. vivax)* has emerged as the primary cause of malaria in Cambodia. Achieving malaria elimination and securing malaria-free certification requires a focused effort on addressing *P. vivax* malaria. This is essential because the elimination of *P. vivax* often lags behind that of *Plasmodium falciparum*, making it a critical component in the overall strategy. This study assesses the feasibility of the Mass Drug Administration (MDA) and *P. vivax* Serological Testing and Treatment (PvSeroTAT) integrated with Reactive Case Detection (RACD) in two of the highest malaria burden operational districts of Cambodia and examines the potential for integrating these two approaches with existing malaria elimination efforts.

**Methods:** This study employs an observational, prospective cohort design. MDA with chloroquine (CQ) will be conducted in Stung Treng through four monthly rounds, while RACD with PvSeroTAT will be implemented in Sen Monorom, targeting households near confirmed *P. vivax* cases. Data on coverage, compliance, cost, and stakeholder perceptions will be collected through surveys, interviews, and malaria case monitoring. A Composite Feasibility Index will integrate quantitative and qualitative indicators. Cost and budget impact analyses will assess scalability for malaria-endemic districts.

**Discussion:** Innovative and targeted public health approaches and tools are necessary to ensure the elimination of the malaria parasite reservoir, including the hidden hypnozoites. While MDA with CQ clears active blood-stage infections leading to immediate reductions in malaria prevalence, PvSeroTAT can detect past exposure to *P. vivax by* using serological markers allowing for targeted treatment of individuals at risk of developing relapsing infections with an 8-aminoquinoline. This helps reduce the parasite reservoir more efficiently. This study will provide insight into operational feasibility, implementation costs, community acceptance, and long-term sustainability. The findings will guide Cambodia’s malaria elimination efforts through improved surveillance and targeted interventions.

**Trial Registration:** OSF Preregistration: https://doi.org/10.17605/OSF.IO/5KZH7, retrospectively registered 15 October 2025.

## Background

Malaria remains a significant public health challenge, with *Plasmodium vivax* (*P. vivax*) posing unique obstacles to control and elimination. Historically, underestimated as a benign infection, there is evidence to show that *P. vivax* can cause severe morbidity and mortality with increased risks of hospitalization and all-cause death, particularly due to repeated relapses occurring before full hematologic recovery (1–3). *P. vivax* is the dominant malaria species in regions where *Plasmodium falciparum* (*P. falciparum*) incidence is declining, particularly in countries progressing toward malaria elimination (4). This trend is evident in Cambodia (5, 6), where malaria incidence has dropped substantially with the total cases declining by 97% from 2020 to 2024 (7). However, the proportion of malaria cases attributed to *P. vivax* increased from 84% in 2019 to 99% in 2024 (7). In 2023, there were 1,303 cases of *P. vivax* reported across 17 provinces. In 2024, 296 *P. vivax* cases had been recorded, whereas cases of *P. falciparum* had almost reached zero (7).

This indicates that even with significant progress*, P. vivax* continues to pose a risk in malaria-endemic regions (8). The persistence of *P. vivax* malaria is particularly pronounced along the borders with Vietnam and Laos (9). Unlike *P. falciparum*, *P. vivax* has a dormant liver stage (hypnozoites), which can reactivate, causing relapses and sustaining transmission in the absence of new infections (10, 11). As a result, *P. vivax* malaria persists. Globally, it has been observed that *P. vivax* malaria elimination consistently lags behind that of *P. falciparum*. In regions like Jiangsu, China (20-year lag), Paraguay (16-year lag), and Armenia (10-year lag) *P. vivax* was responsible for causing malaria long after *P. falciparum* was eliminated (S1 Fig). This challenge with *P. vivax* necessitates innovative strategies to achieve global and national targets of malaria elimination (10, 12). As Cambodia nears the complete elimination of *P. falciparum* malaria (13), urgent and targeted interventions are essential to accelerate the elimination of *P. vivax* malaria and other species of human malaria.

The Cambodian National Center for Parasitology, Entomology, and Malaria Control (CNM) has made significant progress in reducing malaria transmission through vector control and case management strategies (6). Currently, CNM implements several measures such as routine use of Glucose-6-phosphate dehydrogenase (G6PD) testing considering the risk profiles of people to enhance the safety and overall effectiveness of radical cure, 7-day primaquine (PQ) treatment (Total dose – 3.5mg/kg given over 7 days), Targeted Drug Administration, Active Foci Screening, and Intermittent Preventive Treatment for individuals who access forests (14).

Additionally, CNM regularly conducts Reactive Case Detection (RACD), where, upon confirmation of a malaria index case at a healthcare facility, follow-up is carried out in the patient’s home area (15). This involves screening the index case and surrounding households for malaria infection with rapid diagnostic tests and referring any identified cases for appropriate treatment (14, 15).

Despite these efforts, *P. vivax* transmission persists, requiring additional tools and approaches to accelerate elimination. Two primary challenges to *P. vivax* elimination are i) the inability to detect hypnozoites and ii) the relatively low parasite density of infections. No current diagnostic tools can reliably identify dormant liver-stage parasites, meaning a reservoir of infection remains unidentified in the population, making it difficult to ensure the efficacy of radical cure (16). Standard diagnostic methods, including microscopy and rapid diagnostic tests, often fail to detect asymptomatic infections with low parasite density, which constitute a significant proportion of the transmission reservoir (16).

The inability to detect hypnozoites may be resolved with the development of a new standardized *P. vivax* serological assay. This assay identifies recent (6–9 months) *P. vivax* infections, and emerging evidence suggests that it serves as a reliable proxy for hypnozoite carriers, particularly in low-transmission settings where serological exposure markers demonstrate high accuracy (17, 18). *P. vivax* serological testing and treatment (PvSeroTAT) is an approach that involves screening communities or high-risk individuals with this assay and treating sero-positive individuals with effective radical cure. Integrating PvSeroTAT into the routine RACD activities presents an opportunity to enhance the identification and treatment of hypnozoite reservoirs of *P. vivax* malaria. Modelling studies indicate that a single round of PvSeroTAT implementation may reduce *P. vivax* prevalence by up to 25.2%, while multiple rounds, when combined with improved case management strategies, could achieve a reduction of up to 74.1% (19). A recent study with PvSeroTAT has shown exciting promise for its use among highly mobile individuals living within communities in malaria endemic areas in Cambodia (20).

Hard-to-detect, sub-patent infections pose a significant challenge to *P. vivax* elimination, necessitating population-wide interventions such as Mass Drug Administration (MDA) with chloroquine (CQ) (21). The World Health Organization (WHO) currently recommends treating blood-stage CQ-susceptible *P. vivax* infections with CQ or an artemisinin-based combination therapy (ACT), and using ACT in areas with confirmed CQ resistance (22). To clear dormant liver-stage parasites, blood-stage treatment must be paired with an 8-aminoquinoline drug, either primaquine (given as a 14-day regimen of 0.25 - 0.5 mg/kg/day or a 7-day regimen of 0.5 -1 mg/kg/day) or tafenoquine, a single 300 mg dose (22). However, WHO conditionally recommends MDA without an 8-aminoquinoline drug to reduce vivax malaria transmission, and advises against using such drugs alone (i.e. without G6PD testing) in mass treatment campaigns (22).

MDA can be a strategy to rapidly reduce the malaria burden by clearing the asymptomatic and low-density infection reservoir that sustains transmission (19). Previous studies using PQ-based mass-drug strategies, such as Targeted Mass Treatment (TMT) and MDA, have demonstrated their effectiveness in *P. vivax* malaria control (23, 24). CQ, once sidelined because of resistance in malaria parasites, is being reevaluated. New evidence indicates a resurgence in CQ susceptibility of *P. falciparum* in some endemic areas (25–27). For treating *P. vivax,* CQ’s efficacy was confirmed by a study in northeastern Cambodia, where parasites were cleared in all patients by day 3 without evidence of resistance (28). Resistance was ruled out through drug concentration analysis in cases of recurrences that occurred within two months (median 49 days). These findings underscore CQ’s potential in MDA programs to rapidly reduce *P. vivax* reservoirs. MDA with CQ and PQ has previously contributed to malaria elimination in Greece, demonstrating its potential when combined with other control measures (29).

However, within the Cambodian context, it is unknown whether these interventions are feasible to implement at the community level for CNM. MDA may rapidly reduce *P. vivax* transmission, but its effect wanes within 1–3 months (30). Therefore, according to WHO guidelines, MDA should be implemented as part of a broader malaria elimination strategy, including case-based surveillance, parasitological diagnosis, effective treatment for hypnozoites, and appropriate prevention measures to minimize the risk of resurgence after the intervention ends. Thus, with the goals of achieving malaria elimination in 2025 and preparing for WHO certification in 2028/2029 (31), CNM is evaluating two novel strategies: MDA with four rounds of CQ and RACD using PvSeroTAT. These approaches aim to reduce the parasite reservoir and prevent relapses by addressing both blood-stage and liver-stage infections, respectively.

This study will assess the operational feasibility, costs, and acceptability of these two strategies in the highest malaria-burden districts. CNM, in collaboration with the Center for Health and Social Development (HSD) and with support from MMV, will conduct a 14-month observational study to evaluate these interventions. The findings will provide critical evidence to inform Cambodia’s *P. vivax* elimination efforts by 2025. While significant progress has been made in eliminating *P. falciparum* malaria, operational feasibility, and costs of these novel strategies for *P. vivax* elimination remain uncertain, warranting further research and evaluation.

### Study Objectives

#### Primary Objective

- To determine the operational feasibility of RACD with PvSeroTAT radical cure based on G6PD testing, and 4 rounds of MDA with blood-stage treatment only.

#### Secondary Objectives

- To evaluate the cost of RACD with PvSeroTAT (including point-of-care quantitative G6PD test, pregnancy test, and treatment with ACT and PQ for 7 days (3.5mg/kg total dose) or PQ 8 weeks compared to 4 rounds of MDA with blood-stage treatment only (CQ).
- To explore community, healthcare workers, and policy makers’ acceptability and perceptions of CQ MDA and PvSeroTAT.

#### Exploratory objectives

- To assess the effectiveness of MDA with CQ in reducing clinical *P. vivax* malaria cases in the community.
- To assess the effectiveness of RACD with PvSeroTAT using ACT + PQ for 7 days (3.5 mg/kg total dose) and for 8 weeks (based on G6PD status) in reducing clinical *P. vivax* malaria cases in the community.
- To assess the safety of MDA with CQ and RACD with PvSeroTAT using ACT + PQ for 7 days (3.5 mg/kg total dose) and PQ for 8 weeks.

By generating evidence on the operational feasibility and cost implications of these strategies, this study will provide policymakers and malaria control programs in Cambodia and other malaria-endemic regions with valuable insights as they transition towards elimination. If proven successful, the findings may also inform WHO policy guidance on malaria elimination strategies.

## Methods

### Study Design

This study will utilise an observational, prospective cohort design to assess the operational feasibility of public health strategies to eliminate *P. vivax* malaria in Cambodia. CNM, in collaboration with health staff from national public health facilities and with the support of the HSD Team, will implement MDA in Stung Treng and RACD with PvSeroTAT in Sen Monorom.

Participants receiving MDA will be prospectively followed to assess the operational feasibility of this intervention (Fig 1). For PvSeroTAT, previously identified index cases for RACD in 2024 will be revisited to incorporate serological assessments. Additionally, in-depth qualitative interviews will be conducted to explore insights into stakeholder perceptions and to collect bottom-up cost data relevant to the implementation process. Clinical cases of *P. vivax* malaria will be monitored throughout the follow-up period.

**Fig 1.**
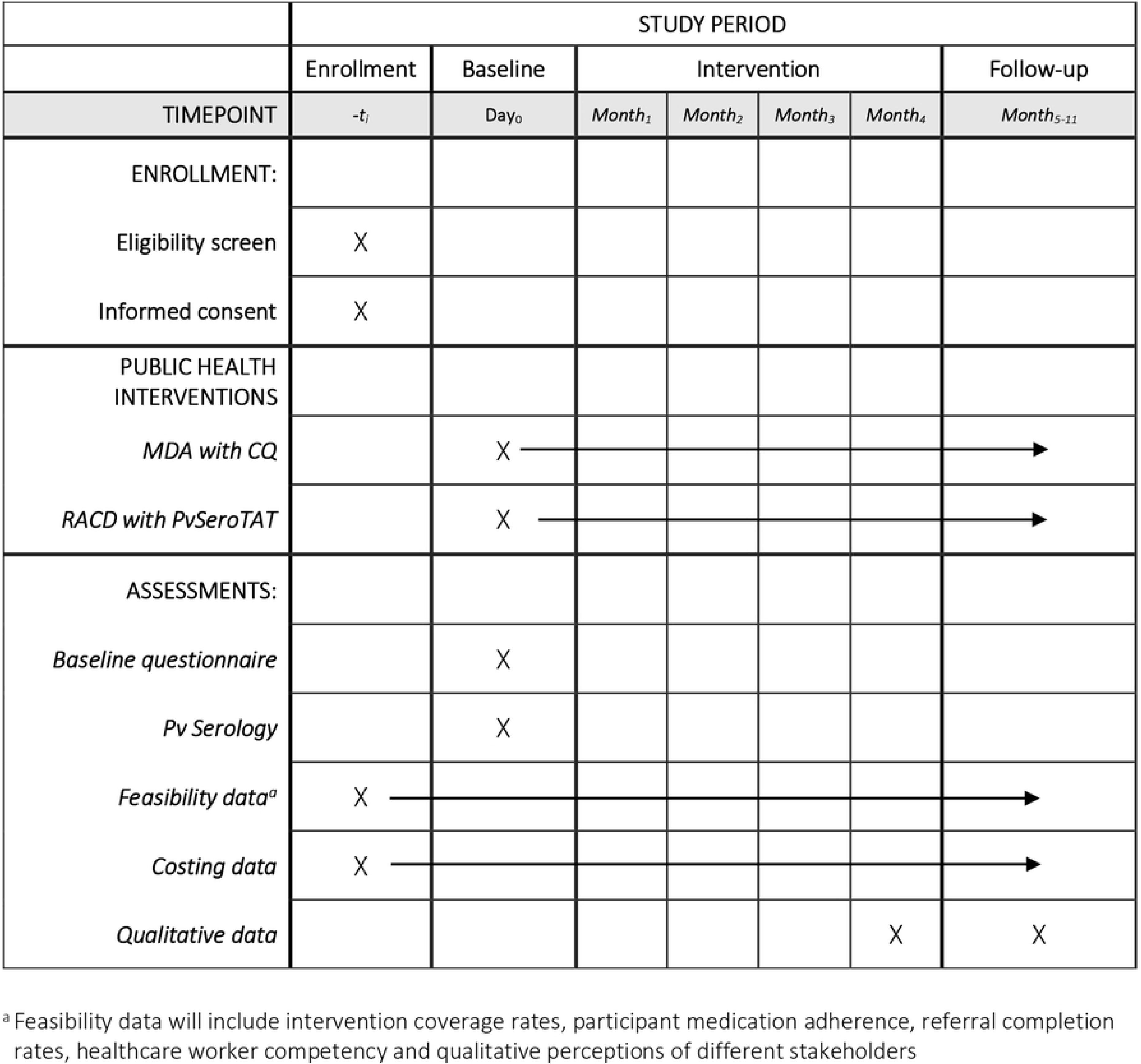
SPIRIT schedule – Overview of study enrolment, interventions and assessments.

### Study Sites

The study will be conducted in two Operational Districts (ODs) in northeastern Cambodia: Stung Treng OD in Stung Treng Province and Sen Monorom OD in Mondulkiri Province. These regions reported the highest caseloads of *P. vivax* malaria in 2023/24 and are comparable in terms of *P. vivax* epidemiology, geography, and population demographics, including age, sex, and migration patterns, including uniform healthcare services. These regions have unique demographic profiles and have been focal points in Cambodia’s malaria elimination efforts due to persistent residual transmission and high-risk populations such as forest goers and mobile workers.

1. Stung Treng: Stung Treng Province, with a population of approximately 159,565 individuals and spanning an area of 11,092 km^2^ of area is bisected by the Mekong River and borders the Lao PDR (Fig 2). Stung Treng consistently ranks as the OD with the highest incidence of malaria in Cambodia in recent years (32, 33). The province also hosts a military base, which is classified as part of a high-risk group for malaria due to increased forest exposure and mobility.
2. Sen Monorom: Located in Mondulkiri Province, Sen Monorom serves as the provincial capital. The province is the largest yet most sparsely populated in Cambodia. With a reported annual parasite incidence of 5.7 cases per 1000 population in 2021, it lies at high risk for malaria (34).

**Fig 2.**
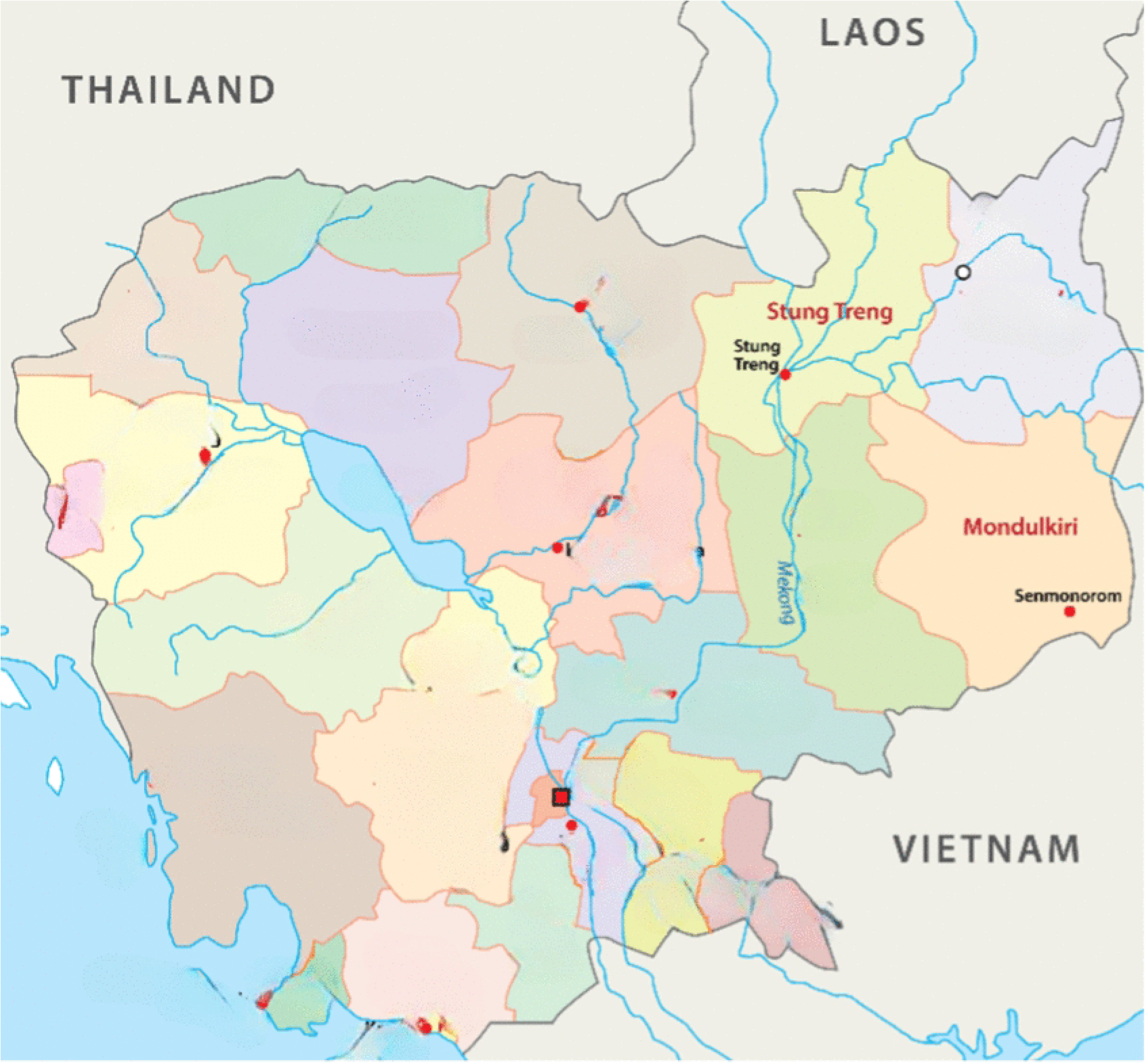
Geographic location of Stung Treng and Sen Monorom OD in Cambodia.

Both regions have experienced demographic shifts due to migration and development activities, influencing their population dynamics and cultural landscapes. In addition, malaria vectors are closely tied to forests in Cambodia and influenced by land use changes. Deforestation, habitat loss, and urbanization alter *Anopheles* vector ecology and sustain malaria transmission (35, 36). These factors, combined with occupational exposures, contribute to the malaria burden in these areas, underscoring the need for targeted elimination strategies.

### Study Sample Size and Population

The estimated sample size for MDA with CQ is 5,000 community people/soldiers (based on the 2022/23 data of the population residing in the active foci in the district) (ref: http://mis.cnm.gov.kh/Dashboard). Soldiers and their relatives in risk areas are included in the MDA target groups.

The estimated sample size for RACD: 6,700 people (assuming four members from 20 households around 98 *P. vivax* cases in the district in the last 9 months, adjusting for 15% absence, non-participation and dropouts) (ref: http://mis.cnm.gov.kh/Dashboard).

### Participant Selection

Participants will be selected based on predefined inclusion and exclusion criteria for MDA and RACD interventions. The criteria are designed to ensure participant safety, adherence to the study protocol, and the validity of the study outcomes.

**Table 1.** Eligibility Criteria For Selection of Participants.

| Criteria | Mass Drug Administration (MDA) | Reactive Case Detection (RACD) |
| --- | --- | --- |
| **Inclusion Criteria** | All community members (adults and children >6 months) living in high-risk areas of Stung Treng OD.<br>Ability to swallow oral medication.<br>Ability and willingness to comply with the study protocol for the duration of the study.<br>Written informed consent from the participant; signed consent from a guardian for children <18 years who provide verbal assent. | Member of a household within 20 households surrounding an identified *P. vivax* malaria case (or within 1 km radius if fewer than 20 houses) in the last 12 months before study initiation.<br>Ability to swallow oral medication.<br>Ability and willingness to comply with the study protocol for the duration of the study.<br>Written informed consent from the participant; signed consent from a guardian for children <18 years who provide verbal assent. |
| **Exclusion Criteria** | Community members (residents and visitors) diagnosed with malaria based on CNM definitions (they will be referred to health centers).<br>History of hypersensitivity reactions or contraindications to chloroquine (CQ). | Pregnant or breastfeeding females.<br>Infants aged <6 months.<br>Participants with a previous adverse reaction or allergy to primaquine (PQ) or the ACT being used.<br>Participants with severe malaria. |

### Recruitment Process

#### MDA in Stung Treng

The recruitment process will target high-risk and hard-to-reach populations, particularly members of the armed forces and forest goers who live and work in remote forested areas along the border. These groups have accounted for the majority of *P. vivax* cases in early 2025. Accordingly, the MDA will be directed at these high-risk groups and their dependents in the Siem Pang district of Stung Treng province. Participants will be required to provide informed consent for a four-round CQ treatment regimen, which will be administered and monitored by trained study personnel. Verbal assent will be taken from children <18 years along with a signed consent from their guardians.

RACD in Mondulkiri: In addition, based on the spatial distribution of *P. vivax* cases in 2024, community members involved in RACD from nine health centers in Sen Monorom will be selected for serological testing. Health center staff will be supported by local authorities, village malaria workers (VMWs), and mobile malaria workers (MMWs) in collecting dried blood spot (DBS) samples from individuals in the target population.

### Interventions

#### Mass Drug Administration (MDA) with Chloroquine (CQ)

MDA will be conducted in Stung Treng using CQ. The intervention will involve four monthly rounds of CQ administration to target blood-stage malaria. CQ will be administered at dosages of 10 mg/kg on days one and two, followed by 5 mg/kg on day three. The administration will be carried out by soldiers, Village Malaria Volunteers and local Health Care Workers through a campaign-style MDA. Participants will receive the first dose of the MDA at central locations, such as soldier camps, community centers, schools, temples, or other designated sites; the day 2 and 3 doses will be given to the participants for intake at home. Oversight for this initiative will be provided by the Provincial Health Department, in conjunction with the CNM and HSD. DBS will be collected from MDA participants during the first round for baseline *P. vivax* serological testing.

#### Reactive Case Detection (RACD) with PvSeroTAT

PvSeroTAT will be implemented among RACD participants from 2024 in Sen Monorom. The strategy involves serological testing of households surrounding an “old” index malaria case (i.e. detected in the last 9 months), within 1 km radius or 20 neighbouring households. DBS collected for serology will be sent to Phnom Penh for analysis. Individuals testing seropositive will be re-identified and referred to facilities for G6PD testing and subsequent treatment with ACT (artemisinin-based combination therapy) and a PQ regimen (either 7 days or 8 weeks, depending on G6PD status) as a *P. vivax* patient in accordance to the national treatment guidelines.

### Data Collection

#### Operational Feasibility Data

Operational feasibility will be evaluated through in-depth interviews (IDIs) with key stakeholders, including the CNM, healthcare workers (HCWs), and other relevant personnel. The interviews will explore compliance, capacity, patient follow-up, safety, training needs, supervision, costs, and logistics.

#### Cost Data Collection

Cost data will be collected through document reviews and Key Informant Interviews (KIIs) at various health system levels (e.g., finance officers, and provincial managers). Structured questionnaires will be administered by project staff to gather retrospective cost data for nine months before the MDA and PvSeroTAT interventions. Costs will be categorized into development, start-up, and recurrent costs, with subcategories including:

- Human resource costs (e.g., healthcare worker allowances for MDA and RACD activities)
- Supervision and travel costs
- Training costs
- Capital costs
- Operating costs (e.g., treatment-related costs, adverse event management)

#### Qualitative Data Collection

IDIs will be conducted with community members, healthcare workers, and policymakers to assess the acceptability and perceptions of RACD with PvSeroTAT and MDA with CQ.

#### MDA and RACD Data Collection

Baseline data on MDA and RACD participants will be collected by trained Cambodian National Health System staff at health facilities. Staff members will use the Malaria Information System (MIS) app to capture data. The MIS app, managed by CNM, will be optimized to include study-specific variables. Participants will be followed-up for medication adherence (self-report) and adverse events (AEs) or severe adverse events (SAEs).

### Data Storage and Security

Data will be collected in both hard copies and digital formats. The MIS app will store data offline and transfer it to a central database once internet access is available. CNM will oversee data storage and security, ensuring confidentiality and restricted access to study personnel only. Personal data will be used strictly for follow-up purposes, while aggregated data will contribute to routine CNM reporting.

### Participant Follow-up

Upon the completion of the MDA, participants will undergo active monitoring every week, conducted either through home visits or telephone communications, for six months. This follow-up will generate valuable data, including the percentage of the population in MDA-targeted areas that received CQ during each of the four months, and new episodes of malaria infection.

Similarly, participants in the RACD will also be followed up weekly for six months after the completion of their treatment. This follow-up will provide insights such as the percentage of seropositive patients that remain contactable after serological analysis and also will include analysis of:

- Malaria recurrence
- Referrals to health centers
- Loss to follow-up due to migration, severe disease, death or other reasons

### Data Management and Analysis

Qualitative Data Management: Qualitative data (audio recordings, interviews and field notes) will be transcribed, translated, and stored in MS Excel for analysis using Nvivo or a similar qualitative software.

Cost Data Management: Cost data will be entered into structured MS Excel sheets and cross-referenced with financial accounts and site surveys. Cost variations between villages or RACD clusters will be analyzed to determine influencing factors.

MDA and RACD Data Management: All MDA and RACD data will be securely stored in paper form and in the MIS. Written consent from all partners (CNM, HSD, MMV) will be obtained before data sharing or publication. To prevent bias, data analysis will be conducted independently by the research team, and only pseudonymized, aggregated results will be provided to non-investigator CNM personnel.

### Data Analysis Plan

To achieve the primary objective of determining the operational feasibility of the two approaches, a Composite Feasibility Index (CFI) will be developed to integrate multiple feasibility indicators into a standardized score. The feasibility of each approach will be assessed by measuring coverage rates, compliance, referral completion, radical cure access, healthcare worker training and competency, supply chain efficiency, and stakeholder engagement. Thematic analysis of in-depth interviews will capture community perceptions, stakeholder acceptability, and operational challenges, ensuring a comprehensive assessment of feasibility. Differences in feasibility perceptions across health administrators, frontline healthcare workers, and community members will be examined using an inductive approach, with qualitative findings incorporated into the CFI through paired comparison analysis or contextual weighting. Table 2 provides key indicators that will be used to calculate CFI along with the types of methods or tools used. Sensitivity analysis will be conducted to test different weighting schemes and validate the robustness of the feasibility index.

**Table 2.** Key metrics or indicators used to calculate CFI.

| Analysis Type | Key Metrics/Indicators | Methods/Tools Used |
| --- | --- | --- |
| Quantitative Analysis | Coverage rates: Proportion of eligible individuals receiving MDA (CQ) and RACD (ACT + PQ). | Descriptive statistics |
|  | Compliance rates: Adherence to the full treatment regimen in both interventions monthly 4-round of CQ and ACT+PQ, via participant interviews. | Descriptive statistics |
|  | Referral completion rates: Number of RACD participants completing follow-up ACT+PQ 7-day (G6PD normal) or ACT+PQ 8-week (G6PD deficient). | Descriptive statistics |
|  | Health worker competency: Post-training assessment scores and compliance with treatment protocols. | Descriptive statistics |
|  | Identification of factors influencing coverage, adherence, and follow-up completion. | Regression modeling |
| Qualitative Analysis | Community perceptions of MDA and RACD acceptability. | Thematic analysis (NVivo) |
|  | Coverage feasibility - explainer | Thematic analysis (NVivo) |
|  | Stakeholder views on operational barriers and facilitators – explainer | Thematic analysis (NVivo) |
|  | Healthcare worker experiences regarding training, supervision, and logistics. – interpret – actual vs their qualitative data | Thematic analysis (NVivo) |
|  | Differences in feasibility perceptions between administrators, healthcare workers, and community members – important for weightage | Inductive approach |

#### Composite Feasibility Index (CFI) Assessment

The CFI will be calculated as a cumulative score integrating quantitative and qualitative feasibility indicators into a standardized value. Quantitative indicators such as coverage and compliance will be weighted more heavily than qualitative factors. The formula for CFI calculation will be:

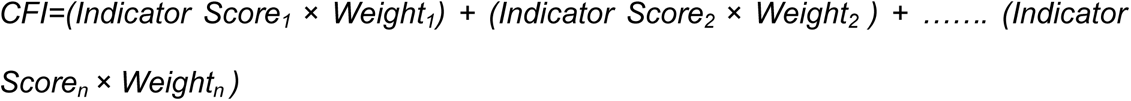

### Cost Analysis

For determining cost, we will use a bottom-up (ingredients-based) costing approach to capture the actual resource utilization for each intervention. Costs will be categorized into development, start-up, and recurrent costs. Initial investment in training materials, protocol development, and system upgrades. One-time expenses such as training healthcare workers, community engagement activities, and procurement of diagnostic kits. Ongoing operational expenses, including personnel salaries, drug supplies, follow-up monitoring, and logistical support. Cost data will be collected through document review and Key Informant Interviews (KIIs) with finance officers, provincial health managers, and implementation staff. Retrospective cost data for the nine months preceding the interventions will be gathered to establish baseline cost estimates. The cost analysis will include the calculation of

- Cost per person per month to estimate the monthly cost of MDA and RACD per individual treated, considering drug procurement, distribution, laboratory costs and personnel expenses.
- Incremental cost per output used to determine the additional cost per person screened and treated in RACD versus the cost per individual covered in MDA.
- Budget Impact Analysis (BIA) to evaluate the financial feasibility of scaling up these interventions to all malaria-endemic Operational Districts in Cambodia.

In addition, cost-consequence analysis will be performed to compare the total costs of MDA and RACD with operational outcomes such as coverage and effectiveness.

### Study Status and Timeline

The study protocol received ethical approval from the National Ethics Committee for Health Research (NECHR) in Cambodia on 26 February 2025 (No. 085 NECHR), with the implementation of the study beginning in March 2025. The study will follow a structured timeline, starting with site preparation and community engagement. Training of study staff and healthcare workers was conducted between March – May 2025. Participant enrolment for MDA and RACD began in April and ended in October 2025. Operational feasibility, costing, and qualitative data collection, accompanied by regular follow-ups will be completed by April 2026. Throughout the study, serological testing, G6PD testing, and radical cure will be administered as needed. The final phases will focus on data analysis, supervision visits, and dissemination of findings through reports and a journal manuscript, with the study concluding in July 2026.

## Discussion

The Cambodian government has set an ambitious target to eliminate all malaria cases by 2025 and is aiming for certification as a malaria-free country in 2028. To achieve this, innovative and targeted public health approaches and tools are necessary to ensure the complete removal of the parasite reservoir. This study is designed to assess the operational feasibility and costs of two public health strategies—MDA with CQ and RACD with PvSeroTAT. Both these strategies present unique operational advantages and challenges in the Cambodian context. MDA with CQ aims to rapidly reduce malaria by administering antimalarial drugs to an entire community, targeting symptomatic and asymptomatic infections. This strategy has been successful in reducing *P. falciparum* prevalence in other near-elimination settings (22), and is now being evaluated for *P. vivax* control. Studies have demonstrated that MDA can lead to immediate reductions in malaria prevalence (29). The sustainability of MDA’s impact is estimated to be influenced by factors such as transmission intensity, coverage rates, and the inclusion of additional control measures. Because WHO does not recommend MDA with PQ for *P. vivax* malaria due to safety concerns, the CNM has prudently opted for CQ, which is conditionally endorsed by the WHO, as a cost-effective alternative (22).

One significant concern with MDA is the potential development of drug resistance, especially if MDA is implemented without adequate coverage or adherence, as observed for *P. falciparum*. However, in Cambodia, *P. falciparum* transmission has been virtually eliminated, with no locally acquired cases reported in recent months (only two cases detected from January to July 2025 were identified as internationally imported), and *P. vivax* now represents the predominant malaria species. While *P. vivax* can also develop resistance to CQ, at least theoretically, if the selective pressure is high enough, this has not been evident in Cambodia. Targeted MDA is warranted to address the remaining reservoir of transmission and accelerate progress toward elimination. Implementing MDA remains logistically demanding, requiring high population coverage, strong community engagement, and sustained adherence. The WHO has provided guidelines to assist national malaria control programs in effectively organizing MDA campaigns, emphasizing the importance of integrating MDA with other malaria control strategies, such as vector control and continuous surveillance, to enhance and sustain its impact (22). In Cambodia’s current context where malaria transmission is increasingly focalized with only a few cases remaining in the country, MDA with CQ offers a potential solution for rapidly reducing the burden of *P. vivax* in endemic areas. Its success, however, hinges on well-coordinated delivery systems and robust follow-up mechanisms.

One major limitation of CQ-based MDA is its inability to target the hypnozoite reservoir in the liver, potentially allowing relapses to sustain transmission. PvSeroTAT represents a novel approach to identifying and treating individuals who may harbor hypnozoites, with many use cases such as outbreak response, preventing reintroduction, risk stratification, and focal screening in high-risk groups (37). Unlike conventional reactive case detection, which only identifies active infections, PvSeroTAT uses serological markers to detect past exposure to *P. vivax* (37). This approach allows for targeted treatment of individuals at risk of developing relapsing infections, which helps reduce the parasite reservoir more efficiently. As widespread administration of PQ (one of the relapse prevention treatments for *P. vivax*) can pose risks to individuals with G6PD deficiency, RACD ensures that only eligible individuals are referred for G6PD testing and receive PQ for either 7 days or 8 weeks. This 8-week treatment strategy mitigates the risk of drug-induced hemolysis in those with G6PD deficiency. It also aligns with WHO recommendations for targeted radical cure strategies and provides a long-term solution to breaking transmission cycles (22). Tafenoquine, the other relapse prevention treatment for *P. vivax*, is not included in this RACD, as it is currently not registered in Cambodia and requires co-administration with CQ for blood-stage treatment (the national guidelines currently use ACT for the blood-stage treatment) (14).

It is important to note that the success of both strategies is highly dependent on community participation and adherence. MDA programs require high coverage and compliance to be effective, yet community acceptability and trust in CQ safety remain potential barriers (30, 38). Previous malaria elimination activities in Cambodia demonstrated that community trust in healthcare providers, proper sensitization, and clear communication improved adherence (39). The study’s approach of engaging VMWs and local healthcare teams is expected to enhance participation and mitigate concerns regarding potential side effects or mistrust in MDA programs.

For RACD with PvSeroTAT, lack of patient adherence to radical-cure treatment is a potential risk. Studies indicate that longer PQ regimens face adherence challenges, particularly in resource-limited settings (40). The integration of community-based follow-ups, phone reminders, and structured supervision in the study design is essential to improving treatment completion rates. Additionally, the study’s qualitative assessments of healthcare workers’ and policymakers’ perceptions will provide insight into factors influencing acceptability and scalability.

Understanding the cost details of MDA vs. RACD with PvSeroTAT is crucial for policy decisions. The study employs a bottom-up costing approach to evaluate direct implementation costs for CNM, considering factors such as drug procurement, laboratory costs, health worker training, supervision, and community mobilization. While MDA may appear more cost-efficient per person treated, the long-term benefits of PvSeroTAT in reducing relapse-associated treatment costs may offer greater sustainability for malaria elimination efforts. However, this comparison will require a cautious interpretation to factor in adjustment of any site-specific factors.

To ensure the study’s effectiveness and adherence to the protocol, multiple monitoring strategies will be employed. Monthly progress reports will document key milestones, challenges, and modifications. Regular site visits will be conducted by the study team to assess site readiness and implementation fidelity. Stakeholder meetings, involving representatives from CNM, MMV, WHO, and other relevant organizations, will be held every month to review study progress and address potential challenges. A digital dashboard will also be implemented for real-time monitoring, allowing researchers to track data entry status and intervention outcomes efficiently.

### Ethical Considerations

Strict confidentiality will be maintained where all the personal data is pseudonymized with an unique number, and the key to link them will be stored separately and securely. Data collection will adhere to ethical principles of lawful, fair, and transparent processing, with explicit use only for tracking patients, treatment adherence, and health monitoring. Access to the data will be restricted to only authorized personnel, and aggregated data will be used for CNM program monitoring. The research team will independently analyze data to avoid bias, with only pseudonymized results shared with non-investigators. Any kind of deviation from the protocol will be documented. All HCWs and VMWs will be responsible for participant safety. Transparency will be ensured by sharing regular updates with participants, communities, and stakeholders. All data collection, processing, and analysis procedures will be transparently reported to ensure scientific validity and unbiased results.

## Conclusion

The success of malaria elimination efforts in Cambodia depends on the implementation of effective, scalable, and cost-efficient strategies. This study will generate crucial evidence on the feasibility, acceptability, and cost implications of MDA and PvSeroTAT. By addressing key operational challenges and identifying best practices, the findings will contribute to Cambodia’s goal of achieving malaria elimination and inform broader regional strategies in the Greater Mekong Subregion. The integration of MDA and PvSeroTAT into CNM’s national strategies has the potential to accelerate the elimination timeline, providing a model for other malaria-endemic countries striving to eradicate *P. vivax*. Both MDA with CQ and RACD with PvSeroTAT hold promise as malaria elimination strategies in Cambodia. MDA offers a broad, rapid reduction of *P. vivax* burden, while RACD with PvSeroTAT targets hypnozoite carriers, preventing long-term transmission. Their success will depend on operational feasibility, adherence rates, costs, and community engagement. The study results will inform CNM and potentially influence global malaria elimination strategies.

## Data Availability

No datasets were generated or analysed during the current study. All relevant data from this study will be made available upon study completion.

## List of abbreviations

ACT: Artemisinin-based Combination Therapy
CNM: National Center for Parasitology, Entomology, and Malaria Control
CQ: Chloroquine
DBS: Dried Blood Spots
G6PD: Glucose-6-phosphate dehydrogenase
HCW: Healthcare worker
HSD: Center for Health and Social Development
MDA: Mass Drug Administration
NECHR: National Ethics Committee for Health Research
OD: Operational District
PQ: Primaquine
PvSeroTAT: Plasmodium vivax Serological Testing and Treatment
RACD: Reactive Case Detection
VMW: Village Malaria Worker
WHO: World Health Organization

## Declarations

### Ethics approval and consent to participate

Before the commencement of this study, ethical approval was obtained from the National Ethics Committee for Health Research (No. 085 NECHR). In addition, written informed consent will be obtained from all participants in Khmer, ensuring full understanding. Verbal assent will be taken from children <18 years along with a signed consent from their guardians.

### Consent for publication

Not applicable

### Availability of data and materials

Data sharing is not applicable to this article as no datasets were generated or analyzed during the current study

## Authors’ contributions

STK, SS, MS and CAL conceived and designed the study, with contributions from JP, IM, LR, TH, TD and EJ. The protocol manuscript was developed by CAL and MS. All authors have reviewed and approved the final manuscript.

## Acknowledgements

The authors would like to thank Dr Rashmi Srivastava for her assistance in drafting this manuscript.

## Supporting Information

**S1 Fig. Lag between P. falciparum and P. vivax elimination across selected countries and regions**

**S1 Table. SPIRIT 2025 checklist of items.**

